# Longitudinal Assessment of Solid Organ Transplant Recipients with SARS-CoV-2 Infection

**DOI:** 10.1101/2024.11.21.24317576

**Authors:** Will Vuyk, Max Bobholz, Isla Emmen, Andrew Lail, Nicolas Minor, Pavan Bhimalli, Jens C. Eickhoff, Hunter J. Ries, Heather Machkovech, Wanting Wei, Andrea Weiler, Alex Richardson, Carson DePagter, Grace VanSleet, Maansi Bhasin, Sarah Kamal, Sydney Wolf, Aanya Virdi, Taylor Bradley, Angela Gifford, Melanie Benito, Alex Shipe, Rana Mohamed, Jeannina Smith, Nancy Wilson, Thomas C. Friedrich, David H. O’Connor, Jacqueline Garonzik-Wang

## Abstract

**Background:** Compared to immunocompetent individuals, those who are immunocompromised, including solid organ transplant (SOT) recipients, have higher SARS-CoV-2-related morbidity and mortality. We determined the duration of SARS-CoV-2 RNA positivity to evaluate viral persistence in SOT recipients.

**Methods:** This study prospectively followed SOT recipients who recently tested positive for SARS-CoV-2. The duration of viral RNA shedding in nasal swabs and stool samples was tracked, and viral genome sequencing was performed where possible. Persistent infection was defined as a positive nucleic acid amplification test (NAAT) for SARS-CoV-2 at 28 days or later after initial infection. This duration was chosen based on the CDC recommendation that immunocompromised individuals isolate for at least 20 days ^1^, compared to 10 days for non-immunocompromised individuals.

**Results:** Of 30 SOT recipients, 12 (40%) had SARS-CoV-2 RNA in nasal swabs or stool 28 or more days after the first positive SARS-CoV-2 test. IC-015 had high viral loads (Ct<30) at 28 days, with continued detection for 54 days.

**Conclusion:** In 12 of 30 SOT subjects, SARS-CoV-2 RNA was detected at or beyond 28 days post-detection (dpd), despite vaccination and antibody and/or antiviral treatment in most participants. Three subjects tested positive for SARS-CoV-2 RNA past 50 dpd. The CDC recommendation for 20 days of isolation may be insufficient for SOT recipients. Viral persistence in the setting of host immune suppression, coupled with exposure to antiviral treatments, raises concern about the selection of unusual viral variants.

**X (twitter) post:** Include Visual Abstract and following text:

Compared to immunocompetent individuals, those who are immunocompromised, including solid organ transplant (SOT) recipients, have higher SARS-CoV-2-related morbidity and mortality. We determined the duration of SARS-CoV-2 RNA positivity to evaluate viral persistence in SOT recipients.

This study prospectively followed SOT recipients who recently tested positive for SARS-CoV-2. The duration of viral RNA shedding in nasal swabs and stool samples was tracked, and viral genome sequencing was performed where possible. Persistent infection was defined as a positive nucleic acid amplification test (NAAT) for SARS-CoV-2 at 28 days or later after initial infection. This duration was chosen based on the CDC recommendation that immunocompromised individuals isolate for at least 20 days ^1^, compared to 10 days for non-immunocompromised individuals.

In 12 of 30 SOT subjects, SARS-CoV-2 RNA was detected at or beyond 28 days post-detection (dpd), despite vaccination and antibody and/or antiviral treatment in most participants. Three subjects tested positive for SARS-CoV-2 RNA past 50 dpd. The CDC recommendation for 20 days of isolation may be insufficient for SOT recipients. Viral persistence in the setting of host immune suppression, coupled with exposure to antiviral treatments, raises concern about the selection of unusual viral variants.

**Visual Abstract:** 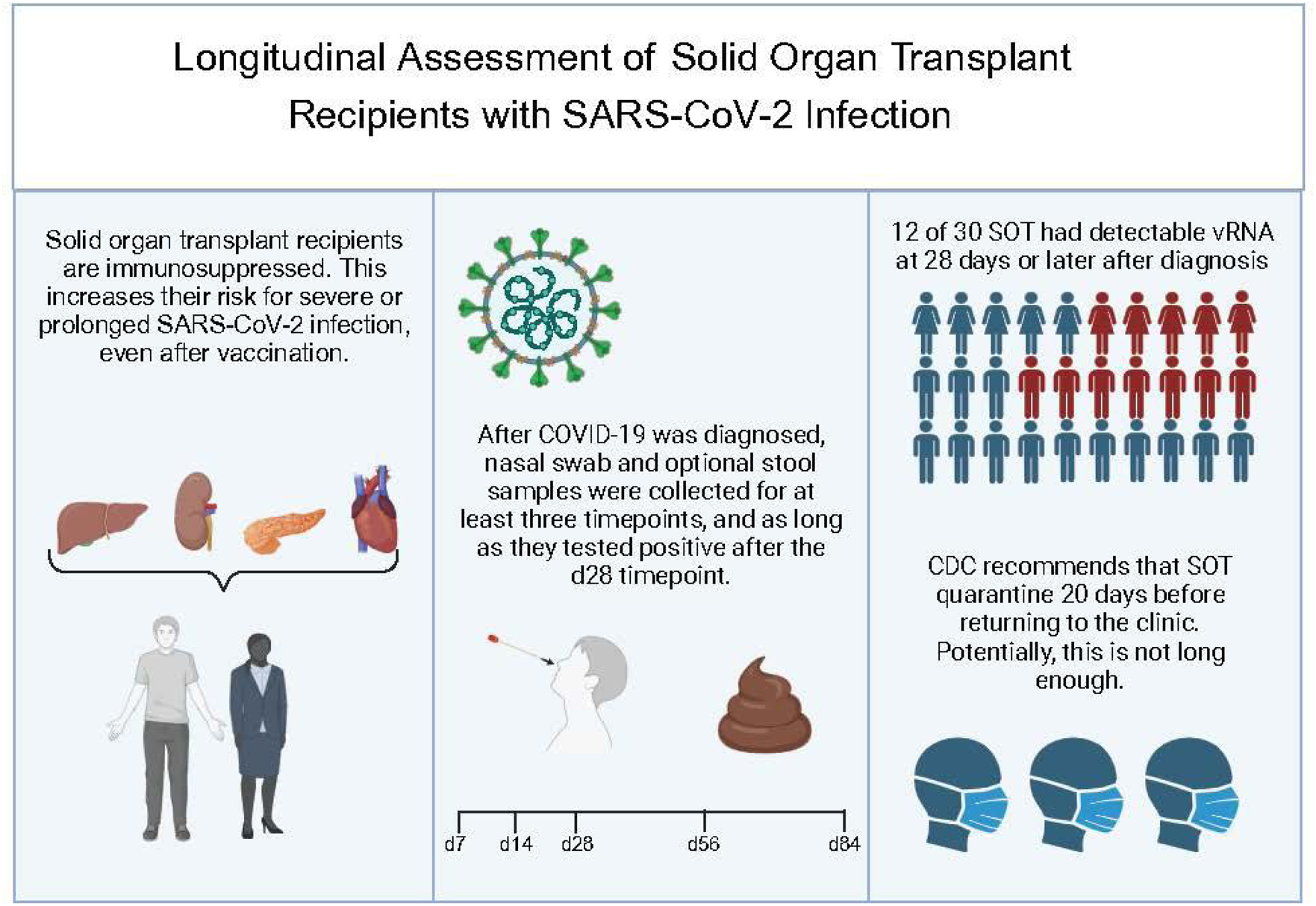

## Introduction

COVID-19 has caused 6.7 million US hospitalizations and 1.1 million deaths as of January 2024 (https://covid.cdc.gov/covid-data-tracker). Immunocompromised (IC) people face higher risks of severe outcomes, especially solid organ transplant (SOT) recipients^2–5^. Although SOT recipients make up 4% of the population, they account for 13% of hospitalizations, 8% of ICU admissions, and 13% of COVID-19 deaths^1,6^. They also face higher risks of death within 28-90 days after infection^7^. While only 0.1% to 0.5% of the general population became persistent COVID shedders, defined as having PCR positive samples taken at least 26 days apart with a Ct<30 where the consensus sequences were of the same lineage^8^, 31 of 59 (53%) of SOT and hematopoietic stem cell transplant recipients became persistent shedders, a 50- to 100-fold higher risk^9^. This risk is highest among people with B cell deficiencies, including hematologic malignancies and uncontrolled HIV infection^9^. Neither vaccination nor subsequent COVID waves have significantly reduced this risk for IC individuals^10,11^.

Typical COVID infections clear within 8-10 days, with median viral clearance at 16 days (average duration 14-17 days)^12,13,14^. Typical COVID infections clear within 8-10 days, with median viral clearance at 16 days. However, some SOT recipients show persistent viral RNA beyond 28 days, with one IC patient’s infection lasting 16 months^14^. These persistent infections raise concerns about new variant emergence, like Omicron^15^.

However, some SOT recipients show persistent viral RNA beyond 28 days, with one IC patient’s infection lasting 16 months. These persistent infections raise concerns about new variant emergence, like Omicron.

## Methods

### Recruitment

We enrolled 40 adult SOT recipients who recently tested positive for COVID-19 (by rapid antigen or PCR). The UW-Madison Health Sciences IRB approved the study (2022-0499). Potential COVID-positive participants were identified by the clinical care team, in both inpatient and outpatient settings, who assessed their interest in participating. Participants were contacted and consented as soon as eligibility was confirmed.

Participants collected nasal swabs, optional stool samples, and blood (if hospitalized). The baseline was defined as the date of SARS-CoV-2 diagnosis or the time of symptom onset, whichever began earlier. Sample collection occurred at baseline (as soon as possible after a positive diagnosis based on either a positive test or symptom onset, followed by a positive test), day 14, day 28, then monthly until both nasal swab and stool (if collected) tested negative, up to 2 years after enrollment.

Participants who joined after day 7 started with the day 14 collection. Except for hospitalized patients, participants self-collected samples at home and shipped them overnight to the lab.

### Patient demographics

Patient demographic data were collected through REDCap surveys and by abstracting patient electronic medical records. De-identified data are presented in Table 1.

**Table 1.**
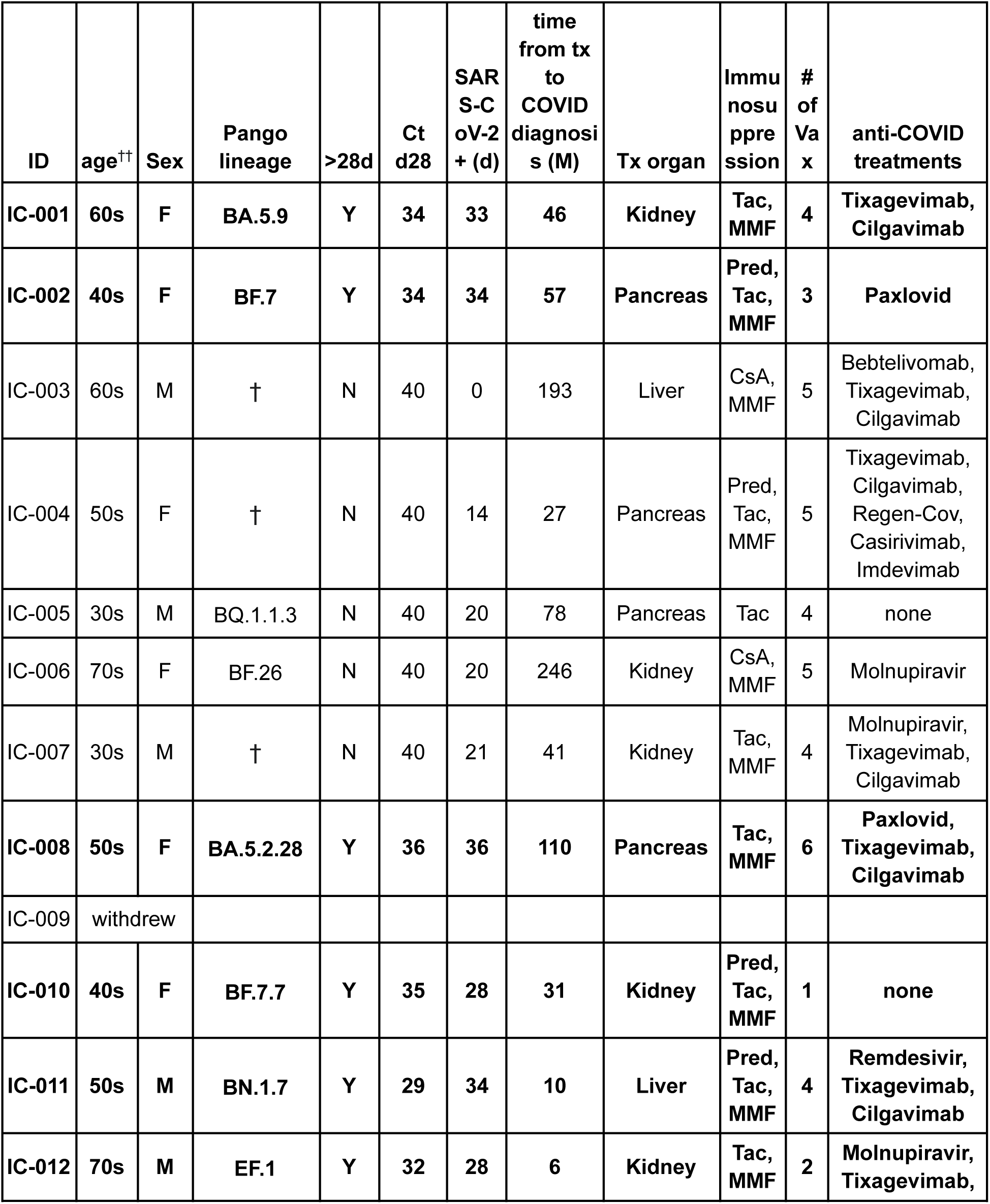
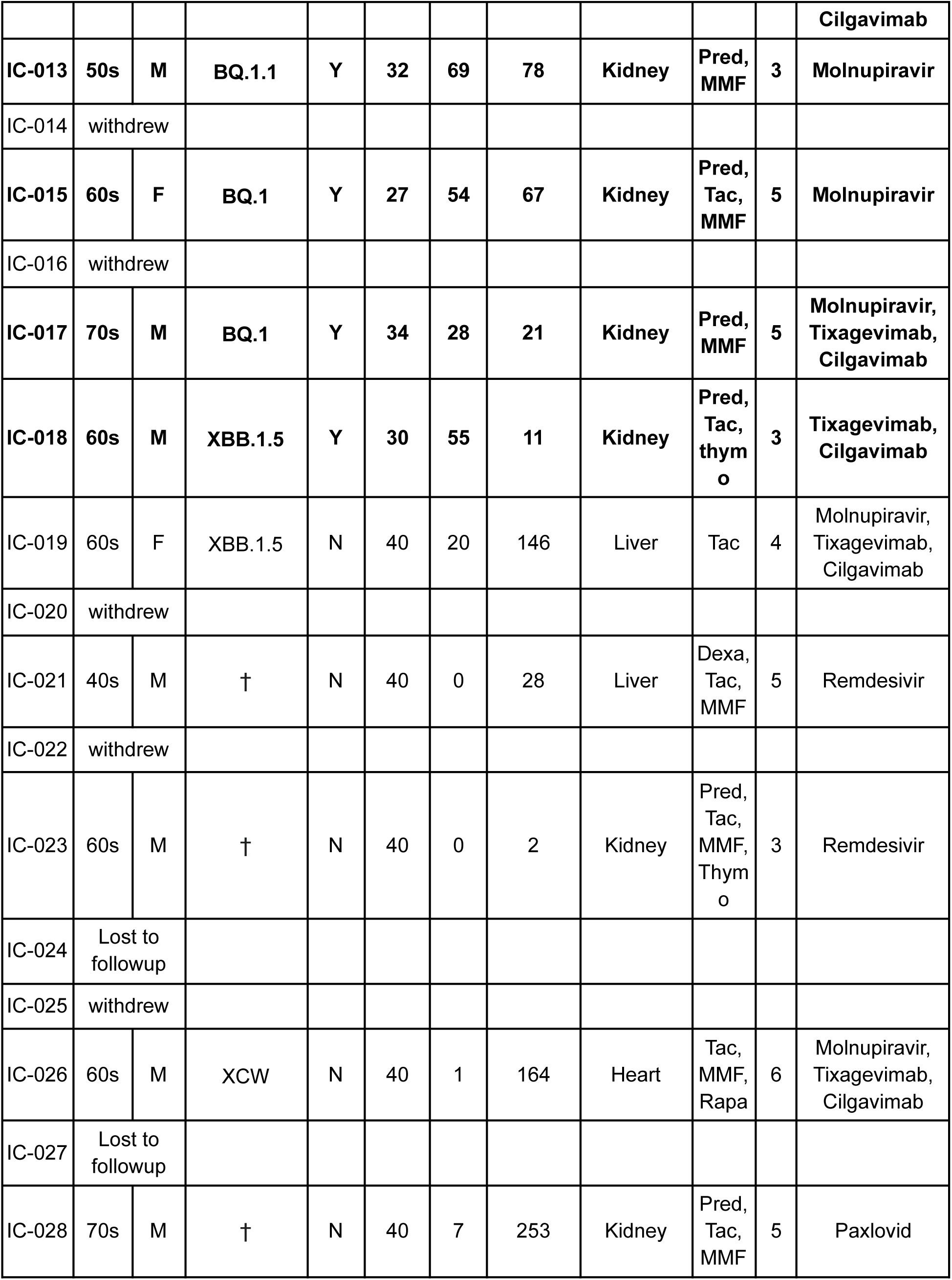
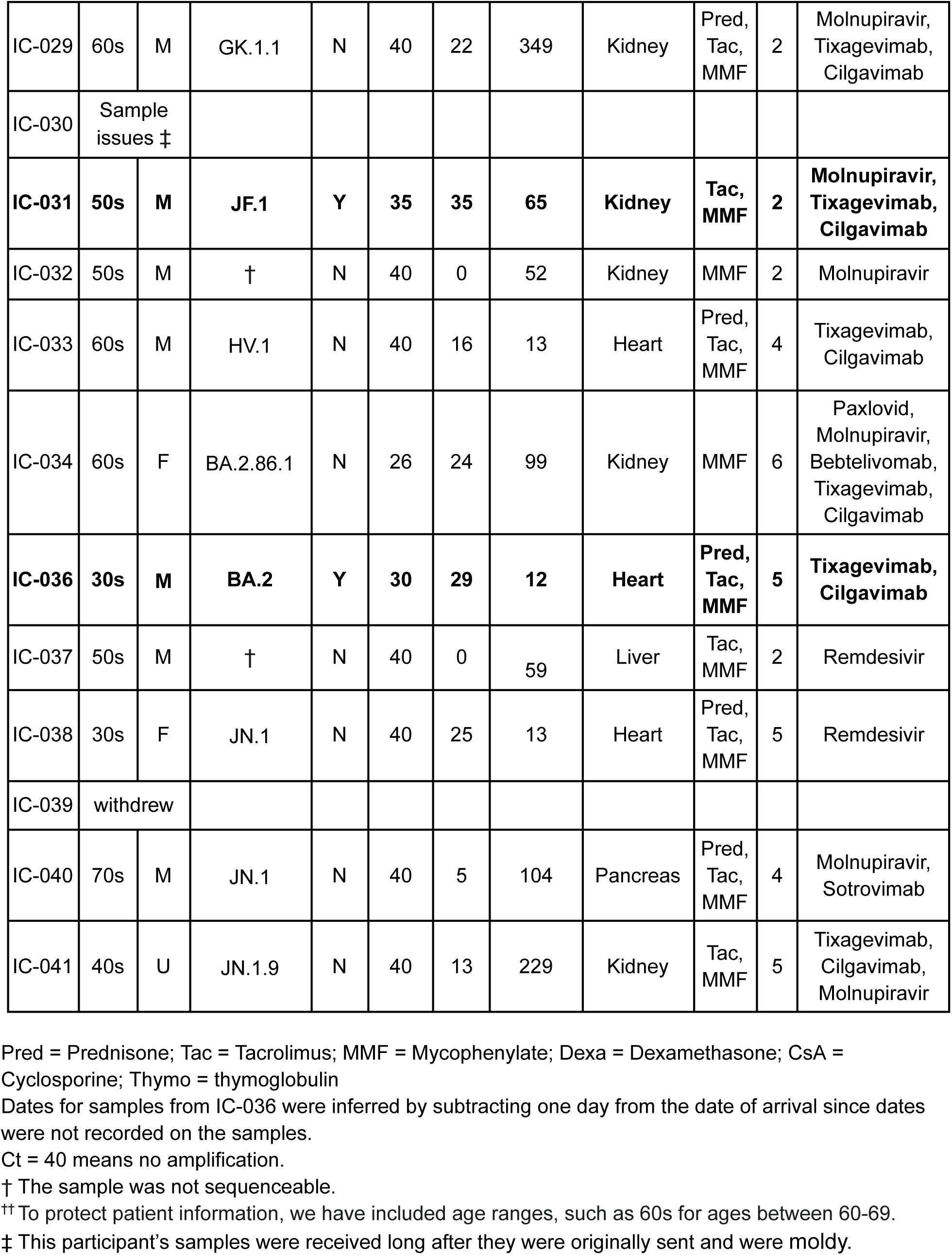
Participant demographics. The bolded participants had detectable SARS-CoV-2 vRNA for at least 28 days after COVID-19 diagnosis. A † in the lineage column indicates that the sample was not sequenceable. The Pango lineages here are from the nasal swab sequencing, because the sequencing was of higher quality in nasal swabs, but Pango assignments from stool samples were the same.

| ID | age<sup>††</sup> | Sex | Pango lineage | >28d | Ct d28 | SAR S-C oV-2 + (d) | time from tx to COVID diagnosis (M) | Tx organ | Immunosuppression | # of Vax | anti-COVID treatments |
| --- | --- | --- | --- | --- | --- | --- | --- | --- | --- | --- | --- |
| IC-001 | 60s | F | BA.5.9 | Y | 34 | 33 | 46 | Kidney | Tac, MMF | 4 | Tixagevimab, Cilgavimab |
| IC-002 | 40s | F | BF.7 | Y | 34 | 34 | 57 | Pancreas | Pred, Tac, MMF | 3 | Paxlovid |
| IC-003 | 60s | M | † | N | 40 | 0 | 193 | Liver | CsA, MMF | 5 | Bebtelivomab, Tixagevimab, Cilgavimab |
| IC-004 | 50s | F | † | N | 40 | 14 | 27 | Pancreas | Pred, Tac, MMF | 5 | Tixagevimab, Cilgavimab, Regen-Cov, Casirivimab, Imdevimab |
| IC-005 | 30s | M | BQ.1.1.3 | N | 40 | 20 | 78 | Pancreas | Tac | 4 | none |
| IC-006 | 70s | F | BF.26 | N | 40 | 20 | 246 | Kidney | CsA, MMF | 5 | Molnupiravir |
| IC-007 | 30s | M | † | N | 40 | 21 | 41 | Kidney | Tac, MMF | 4 | Molnupiravir, Tixagevimab, Cilgavimab |
| IC-008 | 50s | F | BA.5.2.28 | Y | 36 | 36 | 110 | Pancreas | Tac, MMF | 6 | Paxlovid, Tixagevimab, Cilgavimab |
| IC-009 | withdrew |  |  |  |  |  |  |  |  |  |  |
| IC-010 | 40s | F | BF.7.7 | Y | 35 | 28 | 31 | Kidney | Pred, Tac, MMF | 1 | none |
| IC-011 | 50s | M | BN.1.7 | Y | 29 | 34 | 10 | Liver | Pred, Tac, MMF | 4 | Remdesivir, Tixagevimab, Cilgavimab |
| IC-012 | 70s | M | EF.1 | Y | 32 | 28 | 6 | Kidney | Tac, MMF | 2 | Molnupiravir, Tixagevimab, |
|  |  |  |  |  |  |  |  |  |  |  | **Cilgavimab** |
| **IC-013** | **50s** | **M** | **BQ.1.1** | **Y** | **32** | **69** | **78** | **Kidney** | **Pred,**<br>**MMF** | **3** | **Molnupiravir** |
| IC-014 | withdrew |  |  |  |  |  |  |  |  |  |  |
| **IC-015** | **60s** | **F** | **BQ.1** | **Y** | **27** | **54** | **67** | **Kidney** | **Pred,**<br>**Tac,**<br>**MMF** | **5** | **Molnupiravir** |
| IC-016 | withdrew |  |  |  |  |  |  |  |  |  |  |
| **IC-017** | **70s** | **M** | **BQ.1** | **Y** | **34** | **28** | **21** | **Kidney** | **Pred,**<br>**MMF** | **5** | **Molnupiravir,**<br>**Tixagevimab,**<br>**Cilgavimab** |
| **IC-018** | **60s** | **M** | **XBB.1.5** | **Y** | **30** | **55** | **11** | **Kidney** | **Pred,**<br>**Tac,**<br>**thym**<br>**o** | **3** | **Tixagevimab,**<br>**Cilgavimab** |
| IC-019 | 60s | F | XBB.1.5 | N | 40 | 20 | 146 | Liver | Tac | 4 | Molnupiravir,<br>Tixagevimab,<br>Cilgavimab |
| IC-020 | withdrew |  |  |  |  |  |  |  |  |  |  |
| IC-021 | 40s | M | † | N | 40 | 0 | 28 | Liver | Dexa,<br>Tac,<br>MMF | 5 | Remdesivir |
| IC-022 | withdrew |  |  |  |  |  |  |  |  |  |  |
| IC-023 | 60s | M | † | N | 40 | 0 | 2 | Kidney | Pred,<br>Tac,<br>MMF,<br>Thym<br>o | 3 | Remdesivir |
| IC-024 | Lost to<br>followup |  |  |  |  |  |  |  |  |  |  |
| IC-025 | withdrew |  |  |  |  |  |  |  |  |  |  |
| IC-026 | 60s | M | XCW | N | 40 | 1 | 164 | Heart | Tac,<br>MMF,<br>Rapa | 6 | Molnupiravir,<br>Tixagevimab,<br>Cilgavimab |
| IC-027 | Lost to<br>followup |  |  |  |  |  |  |  |  |  |  |
| IC-028 | 70s | M | † | N | 40 | 7 | 253 | Kidney | Pred,<br>Tac,<br>MMF | 5 | Paxlovid |
| IC-029 | 60s | M | GK.1.1 | N | 40 | 22 | 349 | Kidney | Pred,<br>Tac,<br>MMF | 2 | Molnupiravir,<br>Tixagevimab,<br>Cilgavimab |
| IC-030 | Sample<br>issues ‡ |  |  |  |  |  |  |  |  |  |  |
| **IC-031** | **50s** | **M** | **JF.1** | **Y** | **35** | **35** | **65** | **Kidney** | **Tac,**<br>**MMF** | **2** | **Molnupiravir,**<br>**Tixagevimab,**<br>**Cilgavimab** |
| IC-032 | 50s | M | † | N | 40 | 0 | 52 | Kidney | MMF | 2 | Molnupiravir |
| IC-033 | 60s | M | HV.1 | N | 40 | 16 | 13 | Heart | Pred,<br>Tac,<br>MMF | 4 | Tixagevimab,<br>Cilgavimab |
| IC-034 | 60s | F | BA.2.86.1 | N | 26 | 24 | 99 | Kidney | MMF | 6 | Paxlovid,<br>Molnupiravir,<br>Bebtelivomab,<br>Tixagevimab,<br>Cilgavimab |
| **IC-036** | **30s** | **M** | **BA.2** | **Y** | **30** | **29** | **12** | **Heart** | **Pred,**<br>**Tac,**<br>**MMF** | **5** | **Tixagevimab,**<br>**Cilgavimab** |
| IC-037 | 50s | M | † | N | 40 | 0 | 59 | Liver | Tac,<br>MMF | 2 | Remdesivir |
| IC-038 | 30s | F | JN.1 | N | 40 | 25 | 13 | Heart | Pred,<br>Tac,<br>MMF | 5 | Remdesivir |
| IC-039 | withdrew |  |  |  |  |  |  |  |  |  |  |
| IC-040 | 70s | M | JN.1 | N | 40 | 5 | 104 | Pancreas | Pred,<br>Tac,<br>MMF | 4 | Molnupiravir,<br>Sotrovimab |
| IC-041 | 40s | U | JN.1.9 | N | 40 | 13 | 229 | Kidney | Tac,<br>MMF | 5 | Tixagevimab,<br>Cilgavimab,<br>Molnupiravir |

Pred = Prednisone; Tac = Tacrolimus; MMF = Mycophenylate; Dexamethasone; CsA =
Cyclosporine; Thymo = thymoglobulin
Dates for samples from IC-036 were inferred by subtracting one day from the date of arrival since dates were not recorded on the samples.
Ct = 40 means no amplification.
† The sample was not sequenceable.
†† To protect patient information, we have included age ranges, such as 60s for ages between 60-69.
‡ This participant's samples were received long after they were originally sent and were moldy.

### Nasal swab viral isolation and RT-qPCR

Viral RNA load analysis was performed on anterior nares swab samples after they arrived in the laboratory. Total nucleic acids were isolated using the Maxwell HT Viral TNA custom kit (Promega, Fitchburg, WI) on a Kingfisher Apex instrument (Thermo Fisher, Waltham, MA) following the manufacturer’s instructions. Viral RNA quantification was performed using the CDC’s SARS-CoV-2 N1 RT-qPCR assay commercially available from IDT (Coralville, IA) https://www.fda.gov/media/134922/download. The assay was run on a LightCycler 96 (Roche, Indianapolis, IN) using the TaqMan Fast Virus 1-step Master Mix enzyme (ThermoFisher, Waltham, MA). The assay detection limit is estimated to be 325 genome equivalents/ml swab fluid. To determine the vRNA load, samples were interpolated onto a standard curve consisting of serial 10-fold dilutions of *in vitro* transcribed SARS-CoV-2 N gene RNA.

### Stool sample viral isolation and RT-qPCR

Stool samples were self-collected using stool collection kits (Norgen, Thorold, Ontario, Canada). These kits preserve RNA in stool samples for up to 7 days at room temperature. Total nucleic acids were isolated from approximately 300ug of each stool sample using the Promega RSC Fecal Microbiome DNA kit (Promega, Fitchburg, WI). The isolated TNA was then qPCR tested for SARS-CoV-2 N1, and N2 targets using the GoTaq Enviro Wastewater SARS-CoV-2 System (Promega, Fitchburg, WI)^16,17^ using manufacturer’s instructions. Samples were tested in triplicate for each gene target; also for pepper mild mottle virus (PMMoV), a plant virus ubiquitously present in human stool^18,19^. For samples to be considered positive, PMMoV must be detected, and at least one SARS-CoV-2 gene target must be detected in all three technical replicates. If at least one SARS-CoV-2 target was detected in at least one replicate, but the viral target was not detected in all three replicates, the sample was inconclusive.

### SARS-CoV-2 genome sequencing

For samples with Ct <30, total NA from nasal swab samples was isolated without further concentration. For nasal swab samples with Ct ≥30 and stool samples, isolated total nucleic acids (NAs) were treated with TurboDNAse (Invitrogen, Thermofisher Waltham, MA), cleaned/concentrated with/Zymo RNA clean and concentrate kits (Zymo Research, Irvine, CA), reverse transcribed, amplified, and prepared for Illumina sequencing using the QIAseq Direct SARS-CoV kit with Enhancer and Booster add-ons (Qiagen, Germantown, MD). Prepared 8 pM pooled libraries were sequenced on an Illumina Miseq instrument using a v2 2x150 reagent kit (Illumina, San Diego, CA).

FASTQ files were analyzed using the nf-core/viralrecon 2.5 workflow^20^ using the SARS-CoV-2 Wuhan-1 reference genome (MN908947.3). Runs were considered to pass quality control if there was greater than 90% genomic coverage (less than 3000 Ns), a read depth of at least 10X, and negative control coverage was less than 10X reads.

### Data availability

All SRA data are in bioproject PRJNA1096689. Consensus sequencing data:

https://dholk.primate.wisc.edu/project/dho/public/manuscripts/published/Longitudinal%20Assessment%20of%20Solid%20Organ%20Transplant%20Recipients%20with%20SARS-CoV-2%20Infection/begin.view#

## Results

From 28 Nov 2022 to 03 Apr 2024, 40 participants were enrolled in the study with an average inclusion duration of 47 days (range 24 - 89). Ten were excluded for not completing study procedures (n=3) or patient withdrawal (n=7), resulting in 30 participants in the final analysis (Figure 1, Table 1).

**Figure 1.**
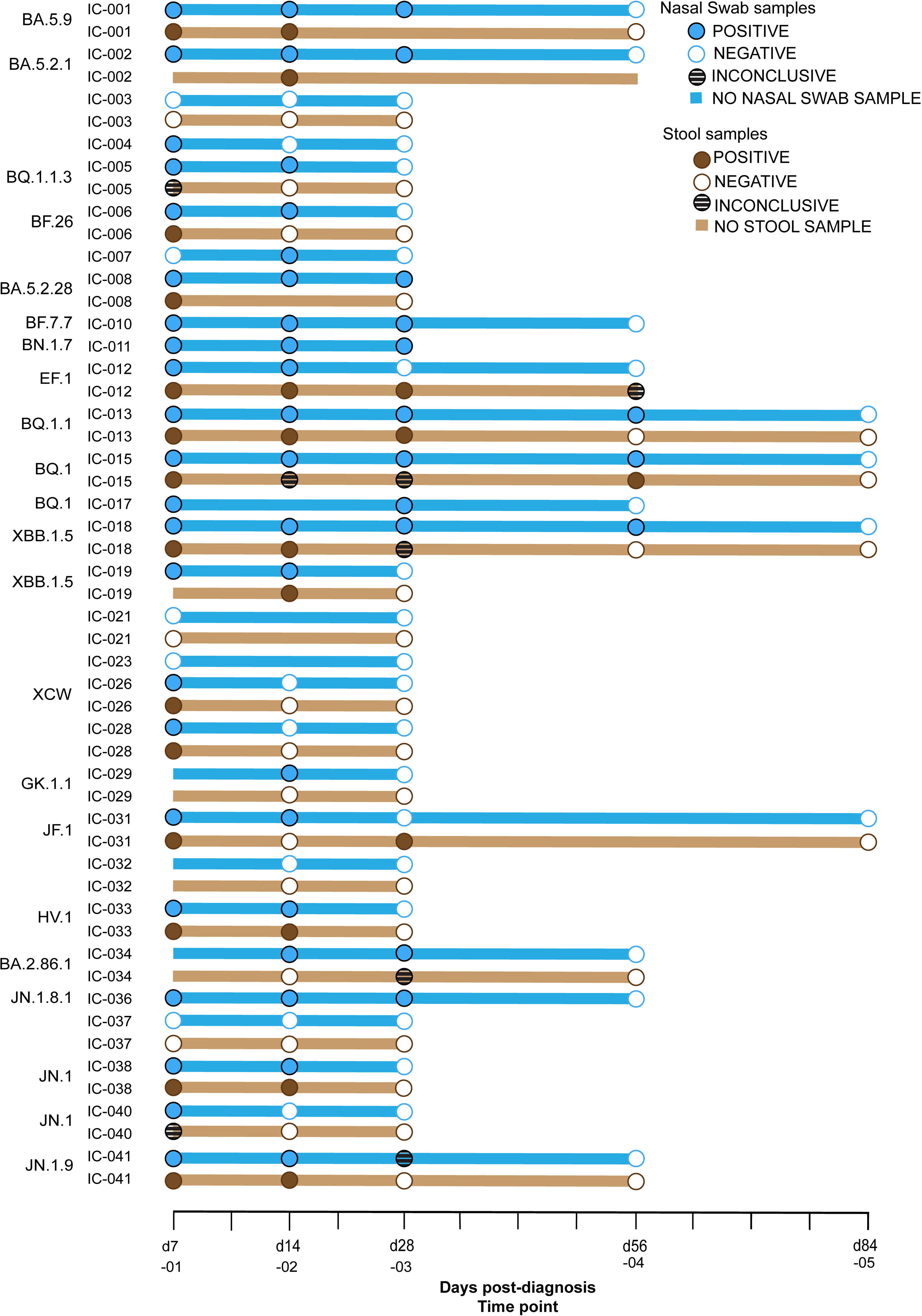
Kinetics of viral RNA positivity in SOT recipients. Filled blue dots signify a **positive** nasal swab. White (with a blue outline) dots indicate a **negative** nasal swab. Dark brown circles indicate **positive** stool samples. White circles (with brown outline) indicate **negative** stool samples. Striped circles are **inconclusive tests**, as defined here: if at least one SARS-CoV-2 target was detected in at least one replicate, but the viral target was not detected in all three replicates, the sample was inconclusive. Stool and nasal swab samples were taken within one day of each other. If sequencing was possible, the Pango lineage is shown to the left of the ID. Participants whose nasal swabs and stool samples both tested negative for SARS-CoV-2 on or before day 28 were not sampled further.

Of these 30 participants, 16 were kidney transplant recipients, 5 were liver transplant recipients, 5 were pancreas transplant recipients, and 4 were heart transplant recipients. The median time from the date of SARS-CoV-2 diagnosis or initial symptoms to the last positive SARS-CoV-2 RT-qPCR test was 23 days (range 1 - 69 days). All participants were vaccinated for SARS-CoV-2 before enrollment, with a median of 4 vaccinations (range 1 - 6 vaccinations). Vaccinations were completed before enrollment, and averaged 12 months before d0, (range 1.5 - 33 M). The median time between transplant and positive SARS-CoV-2 test was 58 months (range 2-349 months). The population was 33% female, with a median age of 60.5 (range 32-79) years. Most subjects received antiviral or antibody therapy upon SARS-CoV-2 diagnosis (Table 1). Six subjects received anti-SARS-CoV-2 monoclonal antibody (MAb) treatment, 10 subjects received antiviral treatment, 12 participants received both MAb and antiviral treatment, and 2 subjects were not treated for COVID-19.

Twelve participants (40%) had detectable SARS-CoV-2 RNA in nasal swabs at day 28 or later, meeting our definition for persistent infection (Figure 1, Table 1). We obtained SARS-CoV-2 sequences from each participant that passed strict criteria of greater than 90% coverage at least 10X depth of coverage, allowing unambiguous SARS-CoV-2 lineage-level classification^21–23^. Three of these 12 subjects had detectable SARS-CoV-2 RNA at 54 days or later (10%) in either nasal swabs, stool, or both. From these participants, we obtained two high-quality SARS-CoV-2 genomes from ≥28 days after either SARS-CoV-2 diagnosis or the first symptoms followed by a positive test. The maximum duration of SARS-CoV-2 RNA positivity was 69 days post-detection (dpd), in a participant whose nasal swab harbored detectable vRNA at day 69.

Twenty-three of the 30 participants also opted to provide stool samples. SARS-CoV-2 RNA was detectable in the stool of four (17%) of these participants at or after day 28 after the first positive test. The maximum duration of SARS-CoV-2 RNA positivity in stool was 54 dpd. Typically nasal swabs had lower Ct than stool samples. Both compartments had the same lineage.

## Discussion

In this prospective study, 12 of 30 (40%) SOT recipients continued to test positive in either nasal swabs or stool at ≥ 28 days, with a maximum of 69 days after the date infection was detected, despite being vaccinated and receiving anti-SARS-CoV-2 mAb and/or antiviral treatment. Additionally, 23 of the participants elected also to send stool samples. Of these 17% tested positive at day 28 or later, out to a maximum of 54 days.

Similar to our results, Raglow et al found that 53% (31 of 59) of SOT or hematopoietic stem cell recipients (HSCT) were RT-PCR positive at day 30 or longer after initial SARS-CoV-2 detection in nasal swabs^24^. Similarly, Li et al describe how viral clearance and evolution are influenced by the type and severity of immunosuppression, with individuals with SOT at lower risk than individuals with uncontrolled HIV or B cell depletion^25^. These results agree with our results and indicate many of these subjects have positive SARS-CoV-2 RNA for longer than the current CDC isolation guidelines for immunosuppressed patients of 20 days^1^. Our results describe an increased prevalence of persistent (>28 days) SARS-CoV-2 infection in transplant patients at 40%, compared to the observed 0.1-0.5% rate in the general population^8^.

Our study is unique in two aspects. Most studies have relied on nasal swab samples, but in a subset of participants, we also collected stool samples. This allowed us to compare viral detection in the respiratory and GI tracts. Additionally, this study was primarily completed remotely. There were a few samples that were collected while the participant was hospitalized, but otherwise, participants were identified by the clinical team, consented remotely, and research kits were shipped to the participants, who collected samples and sent them back. Sample collection in non-clinical settings allowed us to include a greater breadth of participants. Many of our participants lived a significant distance from Madison, so coming to the clinic frequently for sampling would be a burden.

We also did univariate and multivariate analyses to determine whether there were any associations of demographic parameters with persistent infection. While no associations were statistically significant for virus lineage, organ type, immunosuppression, time from transplant was close to being significantly inversely associated with viral persistence with a p value of 0.0525. (Supplemental Tables 1a and b). This must be interpreted cautiously given the participant heterogeneity and relatively small cohort size, but it is possible that given a larger cohort, it would become significant. To determine whether a participant had a persistent infection prior to joining the study, we analyzed whether the SARS-CoV-2 lineages we detected were circulating at the time of diagnosis, indicating recent infection (Supplemental Table 2). SARS-CoV-2 lineages in all participants but IC-026 fell into the expected range of dates where they were circulating in Wisconsin and Illinois, providing no evidence of previously existing persistent infection^26–28^. Typically a difference of 90 days or more would be considered a potential long-term persistent infection^14^.

Among our participants, IC-015 was unique in that we obtained high-quality viral genomes from nasal swabs from this person through d54. However, the stool was negative or inconclusive by d45 therefore, high-quality viral sequence data was not available from stool after day 28 to compare with the nasal swab sequences. Nonsynonymous noncanonical amino acid variants are in the data repository.

The study has several limitations. Our study population was relatively small and consisted exclusively of SOT recipients of the UW Health Transplant Clinic. With few exceptions, patients were diagnosed with SARS-CoV-2 infection using rapid antigen tests at home, which were not sent to us, making it difficult to get samples on the same day as the SARS-CoV-2 diagnosis. However, the first study samples were returned to us with an average of 7.1 d after diagnosis (range 1 - 16), except IC-003, who was a late enrollee. Our sampling strategy was designed to capture samples on days 7, 14, and 28 after diagnosis. It might have been advantageous to sample on day 21, one day after the CDC suggested quarantine period ^1^.

This report adds to accumulating data suggesting that up to 40% of SOT recipients have positive PCR tests for SARS-CoV-2 beyond the 20-day quarantine period^1^ despite treatment with antivirals and anti-SARS-CoV-2 monoclonal antibodies. Persistent infection in SOT recipients could select for lineages that can evade immune responses along with monoclonal antibody and small-molecule antiviral therapies. An improved understanding of the mechanisms and consequences of persistent SARS-CoV-2 infections in SOT recipients will enhance our ability to care for these individuals and address an important potential source of new SARS-CoV-2 variants.

## Abbreviations

CDC: Centers for Disease Control
COVID-19: coronavirus disease 2019
Ct: cycle threshold
dpd: days post detection
IC: immunocompromised
NA: nucleic acid
NAAT: nucleic acid amplification test
PCR: polymerase chain reaction
PPMoV: pepper mild mottle virus
REDCap: research electronic data capture
RNA: ribonucleic acid
RT-qPCR: reverse transcribed quantitative polymerase chain reaction
SARS-CoV-2: severe acute respiratory syndrome coronavirus 1
SOT: solid organ transplant
vRNA: viral RNA

## Acknowledgements

1-Initial funding: Impact of immune failure on SARS-CoV-2 evolutionary potential, CDC Contract Number: 200-2021-11060-FY21 BAA Topic 8 Wisconsin-Madison (Dave and Tom are Co-PIs)

Listed currently on JGW’s IRB

2- DHO / TCF (previously Immune Failure) Discretionary Funds

3 -DHO /TCF 435100-A24-ELCProjE-00 Wisconsin Department of Health Services, ELC Project: E - Enhancing Detection Expansion

4 - JGW Departmental Start Up Funds

## Appendices

**Supplemental Table 1.**
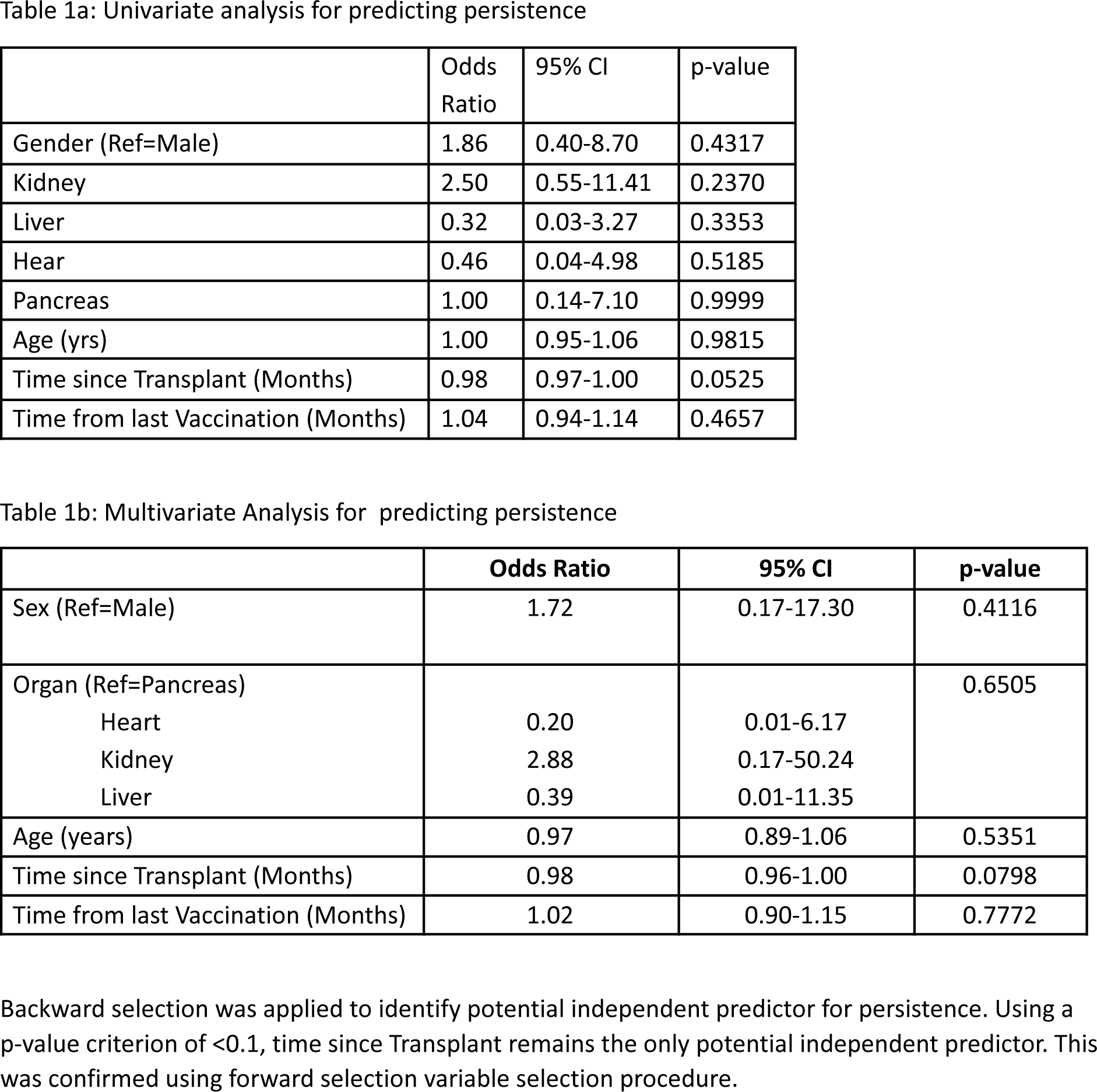
We performed univariate and multivariate analyses to determine whether any demographic parameters were associated with persistent infection. We observed a trend toward significance for the time from transplantation to the likelihood of persistence. The odds of a persistent infection decreased by ∼2% for each month since transplant, after adjusting for organ, age, and sex. Time since transplant for patients with persistent infection was 38.5 months, compared to 88.5 months for those without persistent infection (Odds Ratio 0.98, p-value =0.0525). No other parameters were associated with persistent infection.

**Supplemental Table 2.**
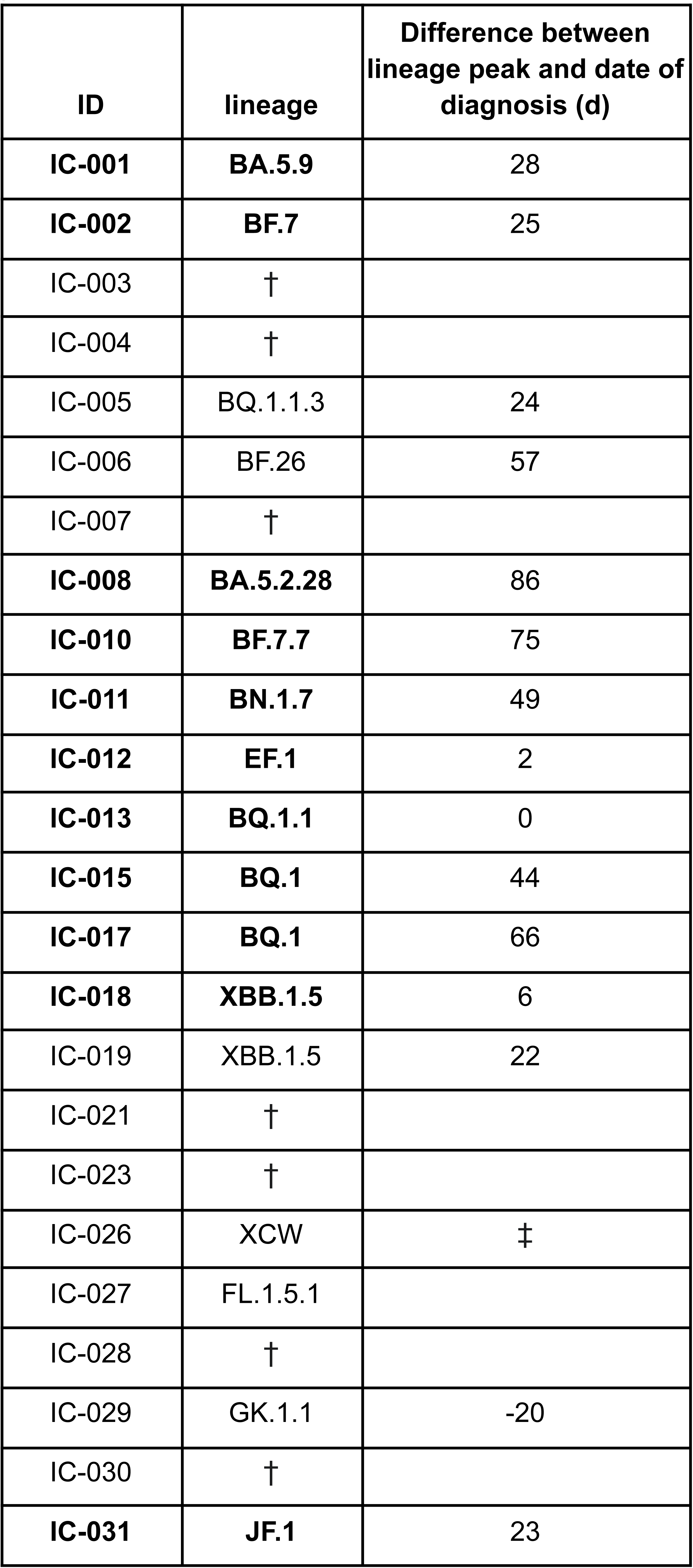
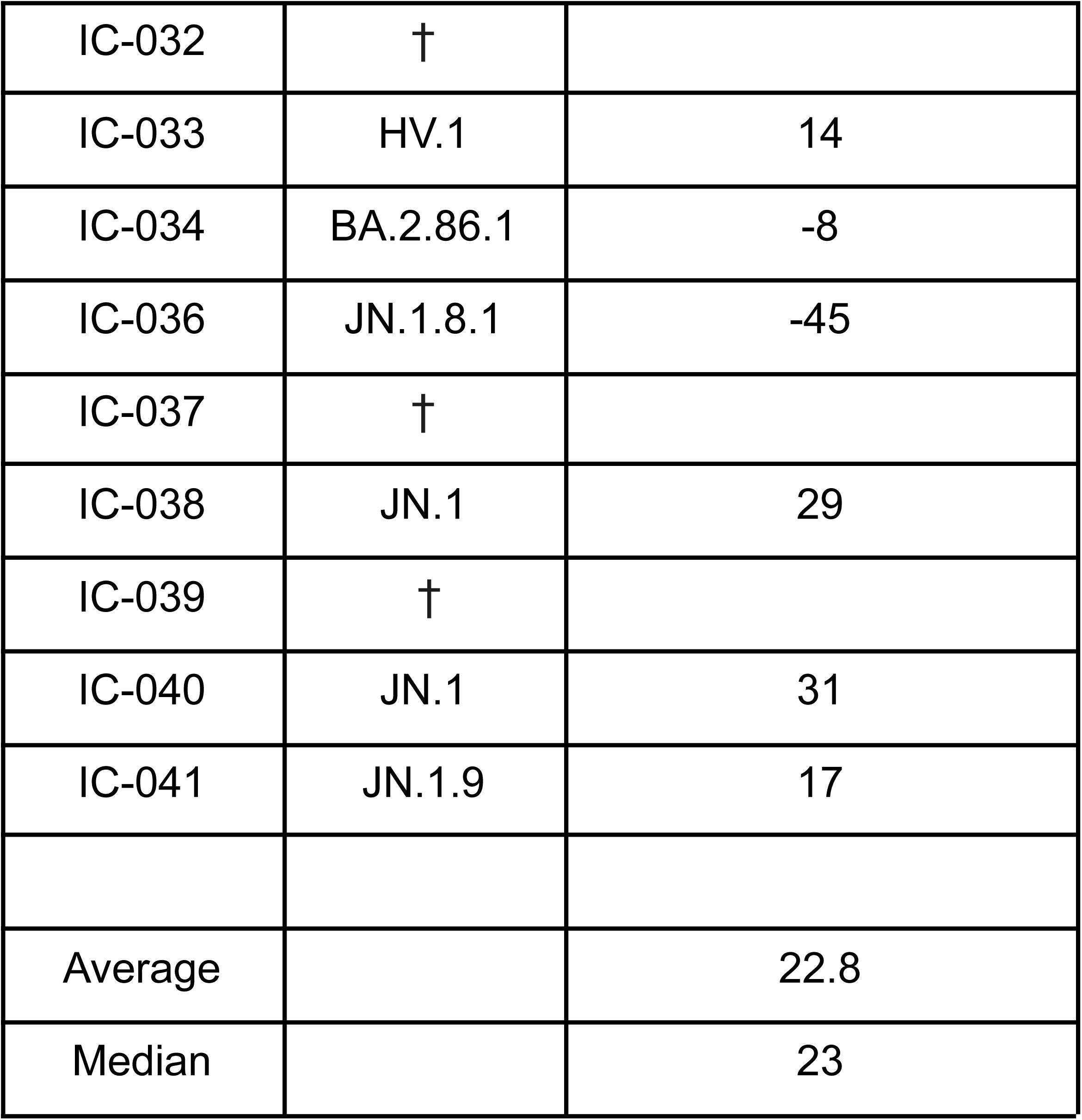
Lineages in study participants occurred within the time period when they were circulating in the Midwest, so it is unlikely that participants had long-term asymptomatic COVID-19 when they entered the study. A † in the lineage column indicates that the sample was not sequenceable. ‡ The variant (XCW) was found at very low prevalence in the US between 7/6/23 and 11/7/23 but was not present in WI.

