## Supplemental Material for "Longitudinal Assessment of Solid Organ Transplant Recipients with SARS-CoV-2 Infection"

### Appendices

**Supplemental Table 1.** We performed univariate and multivariate analyses to determine whether any demographic parameters were associated with persistent infection. We observed a trend toward significance for the time from transplantation to the likelihood of persistence. The odds of a persistent infection decreased by ~2% for each month since transplant, after adjusting for organ, age, and sex. Time since transplant for patients with persistent infection was 38.5 months, compared to 88.5 months for those without persistent infection (Odds Ratio 0.98, p-value =0.0525). No other parameters were associated with persistent infection.

Table 1a: Univariate analysis for predicting persistence

| | Odds Ratio | 95% CI | p-value |
|---|---|---|---|
| Gender (Ref=Male) | 1.86 | 0.40-8.70 | 0.4317 |
| Kidney | 2.50 | 0.55-11.41 | 0.2370 |
| Liver | 0.32 | 0.03-3.27 | 0.3353 |
| Hear | 0.46 | 0.04-4.98 | 0.5185 |
| Pancreas | 1.00 | 0.14-7.10 | 0.9999 |
| Age (yrs) | 1.00 | 0.95-1.06 | 0.9815 |
| Time since Transplant (Months) | 0.98 | 0.97-1.00 | 0.0525 |
| Time from last Vaccination (Months) | 1.04 | 0.94-1.14 | 0.4657 |

Table 1b: Multivariate Analysis for predicting persistence

| | **Odds Ratio** | **95% CI** | **p-value** |
|---|---|---|---|
| Sex (Ref=Male) | 1.72 | 0.17-17.30 | 0.4116 |
| Organ (Ref=Pancreas) | | | 0.6505 |
| Heart | 0.20 | 0.01-6.17 | |
| Kidney | 2.88 | 0.17-50.24 | |
| Liver | 0.39 | 0.01-11.35 | |
| Age (years) | 0.97 | 0.89-1.06 | 0.5351 |
| Time since Transplant (Months) | 0.98 | 0.96-1.00 | 0.0798 |
| Time from last Vaccination (Months) | 1.02 | 0.90-1.15 | 0.7772 |

Backward selection was applied to identify potential independent predictor for persistence. Using a p-value criterion of <0.1, time since Transplant remains the only potential independent predictor. This was confirmed using forward selection variable selection procedure.

**Supplemental Table 2.** Lineages in study participants occurred within the time period when they were circulating in the Midwest, so it is unlikely that participants had long-term asymptomatic COVID-19 when they entered the study. A † in the lineage column indicates that the sample was not sequenceable. ‡ The variant (XCW) was found at very low prevalence in the US between 7/6/23 and 11/7/23 but was not present in WI.

| **ID** | **lineage** | **Difference between lineage peak and date of diagnosis (d)** |
|---|---|---|
| **IC-001** | **BA.5.9** | 28 |
| **IC-002** | **BF.7** | 25 |
| IC-003 | † | |
| IC-004 | † | |
| IC-005 | BQ.1.1.3 | 24 |
| IC-006 | BF.26 | 57 |
| IC-007 | † | |
| **IC-008** | **BA.5.2.28** | 86 |
| **IC-010** | **BF.7.7** | 75 |
| **IC-011** | **BN.1.7** | 49 |
| **IC-012** | **EF.1** | 2 |
| **IC-013** | **BQ.1.1** | 0 |
| **IC-015** | **BQ.1** | 44 |
| **IC-017** | **BQ.1** | 66 |
| **IC-018** | **XBB.1.5** | 6 |
| IC-019 | XBB.1.5 | 22 |
| IC-021 | † | |
| IC-023 | † | |
| IC-026 | XCW | ‡ |
| IC-027 | FL.1.5.1 | |
| IC-028 | † | |
| IC-029 | GK.1.1 | -20 |
| IC-030 | † | |
| **IC-031** | **JF.1** | 23 |

| | | |
|---|---|---|
| IC-032 | † | |
| IC-033 | HV.1 | 14 |
| IC-034 | BA.2.86.1 | -8 |
| IC-036 | JN.1.8.1 | -45 |
| IC-037 | † | |
| IC-038 | JN.1 | 29 |
| IC-039 | † | |
| IC-040 | JN.1 | 31 |
| IC-041 | JN.1.9 | 17 |
| Average | | 22.8 |
| Median | | 23 |
